# Adapting a LAMP Plasmodium vivax Malaria Diagnostic for Use in Low-Resource Settings

**DOI:** 10.1101/2025.04.14.25325264

**Authors:** Anya Chatterjee, Sarayu Parthasarathy, Timothy E. Riedel

## Abstract

Malaria caused by the *P. vivax* parasite can be difficult to diagnose due to dormant phases and genetic variants. Regions such as Sub-Saharan Africa, South America, and Southeast Asia experience diagnostic accessibility issues due to the ‘gold standard’ diagnostic requiring expensive light microscopy of blood samples. This study focuses on expanding the potential of Loop-Mediated Isothermal Amplification (LAMP) as a remote-ready *P. vivax* malaria diagnostic. We detected as few as 10^3^ copies of *P. vivax* 18S rRNA DNA sequence spiked into molecular water and as few as 10^6^ spiked into saliva. These results were reproduced when reactions were incubated in a consumer-ready coffee thermos. This saliva-ready LAMP diagnostic for *P. vivax* shows promise for remote use and increasing diagnostic access to low-resource regions.

## 1. INTRODUCTION

Malaria is an extremely prevalent disease in the 21st century, with up to 500 million new cases and from 1.5 to 2.7 million deaths annually (Bansal et al. 2023). Specifically pervasive in regions such as Central and Eastern Africa, the most vulnerable populations include pregnant women, refugees, and nonimmune travelers (Bansal et al. 2023). While malaria has impacted areas around the world, the “malaria belt,” which includes Sub-Saharan Africa, Southeast Asia, and South America, faces the highest rates of infection, often primarily affecting youth. In Sub-Saharan African countries that are endemic to malaria, many communities in these countries are impoverished due to low economic growth (Gallup and Sachs 2001). Therefore, with improved accessibility for testing, there should be a significant decrease in malaria’s impact on Sub-Saharan Africa, Southeast Asia, and South America.

The five plasmodium species that cause malaria in humans are *P. falciparum, P. vivax, P. ovale, P. malariae*, and *P. knowlesi*, and microscopists determine which plasmodium species a patient is infected with by assessing the unique morphological characteristics of the stained parasitic species in the patient’s blood sample (Oyegoke et al. 2022). Microscopy is the current “gold standard” diagnostic for malaria and is performed after a patient receives a clinical diagnosis based on their symptoms (Oyegoke et al. 2022). The accuracy of microscopy for a malaria diagnosis is high, but it is also expensive due to the need for high-quality microscopes, an experienced technician, reagents, slides, etc., to ensure accuracy in results (Oyegoke et al. 2022).

A malaria diagnostic utilizing the DNA amplification technique Loop-Mediated Isothermal Amplification (LAMP) offers a time-efficient, highly specific, and simple-to-interpret alternative (Notomi et al. 2000). PCR-based diagnostics, though highly accurate, are more expensive than LAMP due to the specialized thermocycling equipment and expertise required to interpret the results. PCR tests also take longer, yielding results in one to a few days, whereas LAMP produces results in about 30 minutes (Soroka and Rymaszewska 2021). While rapid diagnostic tests such as lateral flow test strips are faster than LAMP, they are not as sensitive as molecular diagnostics like LAMP and PCR (Mens et al. 2007).

While *P. falciparum* is the most prevalent and acute form of malaria (Neafsey et al. 2012), this study focuses on *P. vivax*, which commonly causes severe forms of malaria in humans and is twice as genetically diverse as *P. falciparum. P. vivax’s* ability to mutate and give rise to new strains is attributed to its significant genetic diversity (Neafsey et al. 2012). Also, as *P. vivax* and *P. ovale* are the two malaria-causing parasitic species that have dormant stages in the human liver, this can cause *P. vivax* to not circulate as significantly in blood and potentially be missed through microscopic diagnosis (Hulden and Hulden 2011). Prior studies have shown that PCR, LAMP, and other nucleic acid amplification techniques have higher accuracy in detecting the DNA of Plasmodium species in saliva compared to urine (Chai and Chua 2022). LAMP has been proven as a useful diagnostic for malaria in multiple clinical settings in Thailand (Sirichaisinthop et al. 2011, Han et al. 2007).

Given the potential benefits of LAMP testing, we optimized the Han et al. (2007) *P. vivax* LAMP assay that targets the 18S rRNA DNA sequence (18S). We adapted the assay to a simple colorimetric readout and a portable incubator to accept the direct input of saliva. These adaptations to the diagnostic should yield a highly sensitive, affordable, quick, easy to interpret, *P. vivax* malaria diagnostic tool beneficial to malaria-endemic areas.

## 2. MATERIALS AND METHODS

### 2.1 Reaction Mix Preparation

For all LAMP experiments, typical contamination mitigation procedures were followed, including work surfaces wiped with 10% bleach and then 70% ethanol. Negative control reactions were prepared outside of any containment hood and sealed before any associated positive control supplies were thawed. Positive control reactions were prepared in an AirClean600 DNA hood (AirClean Systems, Creedmoor, NC).

All LAMP reactions reported in this study utilized WarmStart Colorimetric LAMP 2X Master Mix (New England BioLabs, Ipswich MA, USA, cat. # M1800L). The NEB 2X Master Mix was combined with 2.5 µL of a 10X primer mix and 9 µL of molecular water for a volume of 24 µL. Three µL of the 18S sequence at known concentrations were added to the positive control samples (PTCs). In contrast, the negative control samples (NTCs) were sealed immediately to prevent contamination. The resulting 24 µL NTC and 27 µL PTC reactions were vortexed briefly to ensure a well-mixed reaction.

For three out of the four PTCs, the 18S sequence was diluted in saliva rather than molecular water. Saliva was treated and buffered using the Lalli et al. 2020 study as a guide. To prepare the saliva, saliva samples were collected from multiple individuals fasting for at least 30 minutes, pooled, and then heat-treated at 65° C for 15 minutes in the heat block. These samples were allowed to passively cool to room temperature, and then buffered 2:1 V/V and vortexed with PBS (900:450 μL for PTCs, 1000:500 μL for NTCs). This treated saliva was spiked with 18S sequence (or not for NTCs), and then 3 μL was transferred to LAMP reactions.

### 2.2 Diagnostic Scoring and Data Collection

The NEB WarmStart Colorimetric LAMP 2X Master Mix contains a pH-sensitive dye, which changes color from pink to orange or yellow upon the amplification of a nucleic acid sequence. Reactions were scored every 10 minutes during incubation by removing the tubes (<15 seconds) from the incubator, photographing them, and quickly returning them to the incubator. Reaction colors were scored with the unaided eye as either pink (P), orange (O), yellow (Y), or with color combinations to represent intermediate levels when needed (e.g., O/Y for an orange-yellow mixture).

### 2.3 Primers and the P. *vivax* 18S Sequence

To detect the prevalence of *P. vivax* using LAMP, a set of primers (Han et al. 2007, Table 1) that targets a segment of the 18S rRNA gene in *P. vivax* was ordered from Integrated DNA Technologies (IDT). The DNA sequences of the primers and 18S DNA sequence were confirmed with Primer-BLAST (NCBI 2024) before ordering them to confirm that the primers annealed to the sequence.

**Table 1.**
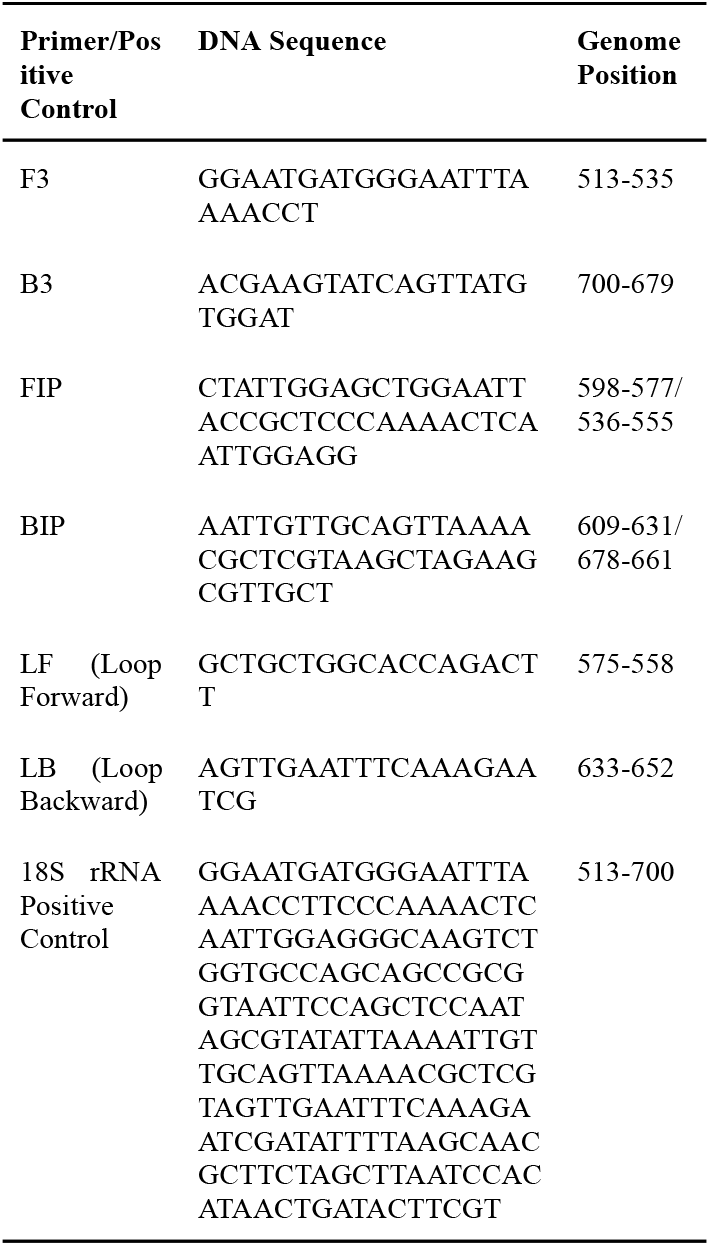
*P. vivax* LAMP primers and positive control 18S sequence (Han et al., 2007) used in this study are reported in the 5’ to 3’ direction.

### 2.4 Primer Mix Preparation and 18S Sequence Dilutions

For each primer (F3, B3, FIP, BIP, FLP, BLP), molecular-grade water was added to make a storage concentration of 100 μM. A 1000 µL 10X working stock of Han primer mix was created according to the concentrations reported in Table 2. After preparing the 10X primer working stock, 330 μL was aliquoted into three 1.5 mL tubes for storage.

**Table 2.** The concentration of each LAMP primer in a 10X stock of Han primer mix. The 1X reaction concentration is the concentration of each LAMP primer in 2.5 μL of the 10X stock of Han primer mix.

| Primer | 10X Stock ($\mu\text{M}$) | 1X Reaction Concentration ($\mu\text{M}$) |
| --- | --- | --- |
| F3 | 2 | 0.2 |
| B3 | 2 | 0.2 |
| FIP | 16 | 1.6 |
| BIP | 16 | 1.6 |
| Loop F (FLP) | 4 | 0.4 |
| Loop B (BLP) | 4 | 0.4 |

Spikes of 18S sequence were prepared by ordering the 18S DNA (IDT gBlock), resuspending it with molecular water, and storing it at a concentration of 10^9^ copies/µL. Specific concentrations of 18S spikes were prepared fresh each day that LAMP was performed via serial dilution with molecular water. LAMP reactions were spiked with 3 μL of either 10^9^, 10^7^, 10^5^, or 10^3^ copies/μL of the 18S sequence in a molecular water or treated saliva matrix (Section 2.1).

3 µL of 1500 µL of treated saliva, which does not have the 18S sequence diluted, is added to three out of the four negative control reaction tubes. These represent the negative control reactions where saliva is the testing matrix. In one of the negative control reaction tubes, no treated saliva is added, so it just includes the reaction mix.

### 2.6 Flask Setup for Remote Testing

For flask-based experiments, an insulated vacuum flask was used to contain the 65° C water and reaction tubes for the LAMP reaction instead of a water bath (Figure 1). For this setup, a vacuum-insulated 750 mL compact stainless steel beverage bottle (Thermos^®^ Bottle, cat. # FBB750SSRT4) was filled with 830 mL of tap water and then preheated to 65° C. A circular tube rack, which was made by cutting and folding aluminum foil, sits on the top of the flask and allows the tubes to be submerged in the water. Eight holes, each the diameter of a LAMP tube, were cut into the rack to hold the LAMP tubes. The temperature of the water contained in the vacuum flask drops 2.6° C over 70 minutes (Figure 2).

**Figure 1.**
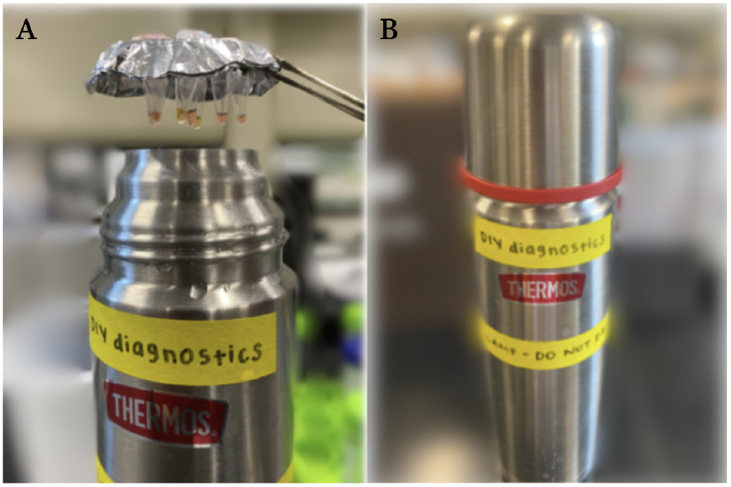
The remote LAMP setup includes the vacuum flask with the lid and the aluminum foil rack holding the reaction tubes. For LAMP reactions, the tube rack made of aluminum foil holds eight reaction tubes and fits on the top of the vacuum flask, which is filled with water at 65° C (A). The lid is then placed on the vacuum flask so that the water temperature stays at around 65° C for 60 minutes (B).

**Figure 2.**
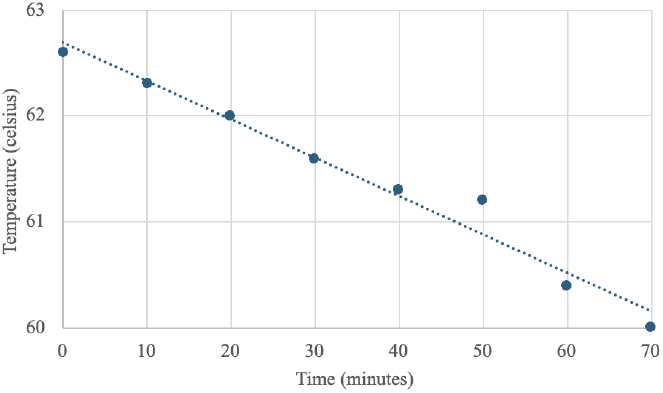
Temperature decline over time in a remote lamp water experiment using a vacuum flask filled with tap water. The graph represents the temperature decrease in tap water over 70 minutes within a vacuum flask in a remote LAMP setup, with an *R*^2^ value of 0.9728. The initial temperature starts at 62.6°C and gradually declines to 60°C by the 70-minute mark.

## 3. RESULTS

### 3.1 Exploring the Sensitivity of the 18S Sequence in a Molecular H_2_O Matrix

To characterize the timing of amplification at different positive control spike concentrations, we challenged the assay against 10^9^, 10^7^, 10^5^, 10^3,^ and 10^2^ copies of the 18 sequence/μL of molecular H_2_O (Figure 3, Table 3-4, Data not shown for 10^9^ and 10^7^ concentrations). When monitored at 10-minute intervals, all concentrations of positive controls turned yellow before 40 minutes except for 10^2^ copies of the 18S sequence/μL of molecular water. All negative controls in the molecular water matrix did not turn yellow until after 50 minutes. Positive controls with 10^2^ copies of the 18S sequence/μL of molecular water showed inconsistent amplification because some of the positive control samples stayed pink. Only one of the positive controls turned yellow before 40 minutes, and the negative controls turned yellow after 50 minutes (Table 5).

**Table 3.**
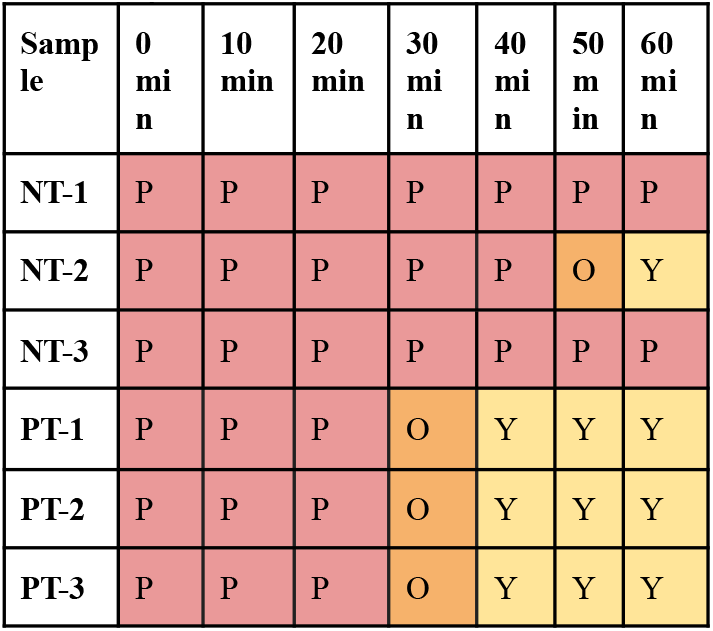
Color scoring results from water bath LAMP reactions. NT samples were spiked with molecular water while PT samples were spiked with 10^5^ 18S sequence.

| Sample | 0 min | 10 min | 20 min | 30 min | 40 min | 50 min | 60 min |
| --- | --- | --- | --- | --- | --- | --- | --- |
| NT-1 | P | P | P | P | P | P | P |
| NT-2 | P | P | P | P | P | O | Y |
| NT-3 | P | P | P | P | P | P | P |
| PT-1 | P | P | P | O | Y | Y | Y |
| PT-2 | P | P | P | O | Y | Y | Y |
| PT-3 | P | P | P | O | Y | Y | Y |

**Table 4.**
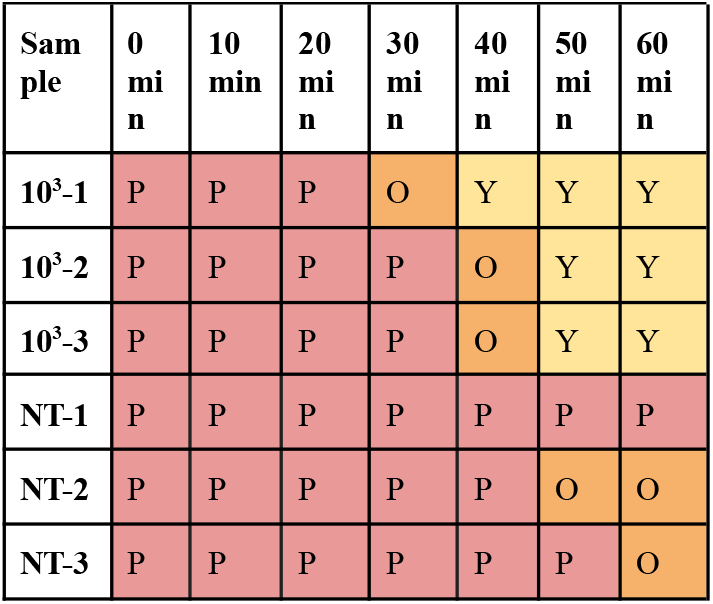
Amplification results with 10^3^ copies/µL 18S sequence resuspended in molecular water. Every 10 minutes in a vacuum flask filled with water at 65° C, the color of the solution in each reaction tube is scored as P (no amplification), O (some amplification), or Y (full amplification).

**Table 5.**
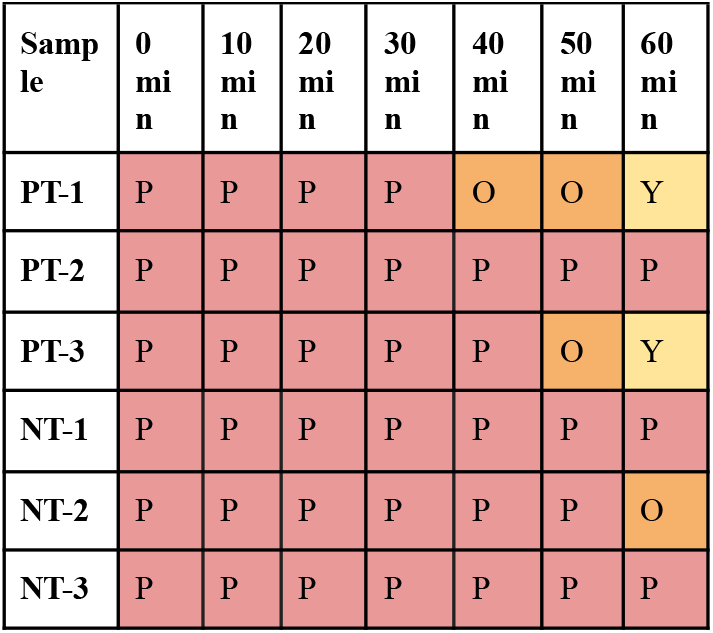
Amplification results with 10^2^ copies/µL 18S spike resuspended in molecular water. Every 10 minutes in a vacuum flask filled with water at 65° C, the color of the solution in each reaction tube is scored as P (no amplification), O (some amplification), or Y (full amplification).

**Figure 3.**
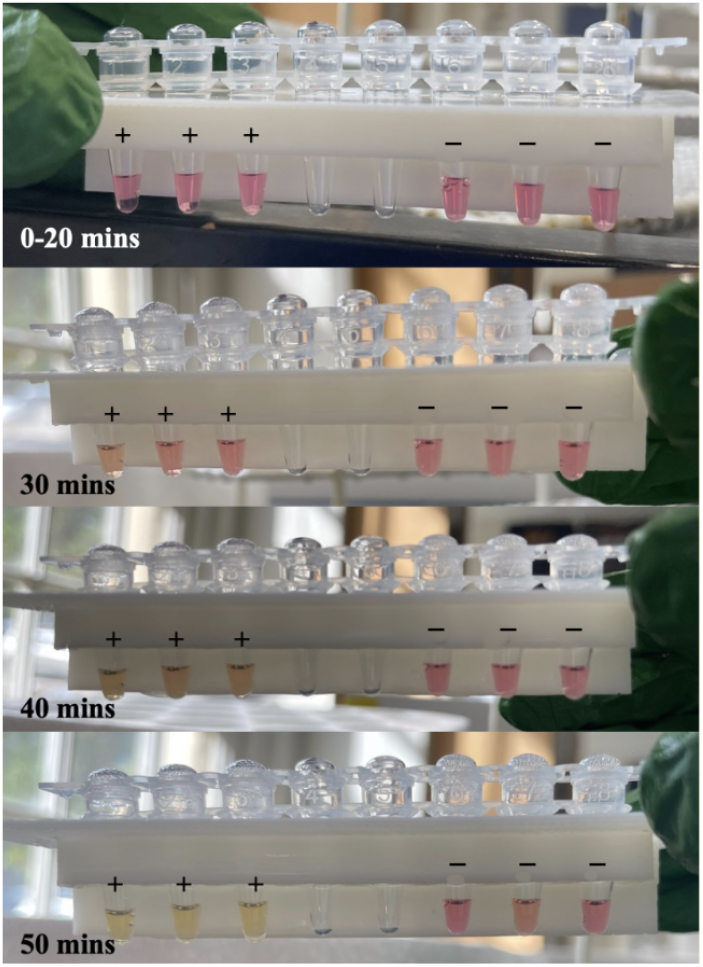
Positive control reaction tubes with 10^3^ copies/μL of 18S spike resuspended in molecular water versus negative controls. All tubes were incubated in a lab-grade water bath. Tubes labeled with a plus sign were spiked with 3 μL of *P. vivax* 18S rRNA sequence. Tubes labeled with a negative sign were spiked with 3 μL of molecular water. Tubes were photographed and scored as pink, orange, or yellow in 10-minute increments.

### 3.2 Testing Assay in a Saliva Matrix and Vacuum Flask

With LAMP reactions on positive controls with 10^9^ and 10^7^ copies of the 18S sequence/μL of saliva, the solutions changed color from pink to orange at 30 minutes. If a color change occurred at 30 minutes, this means that the Han primers were specifically bound to the 18S rRNA sequence, and the 18S sequence amplified between 20 to 30 minutes. With a LAMP reaction on four replicate positive controls with 10^6^ copies of the 18S sequence/μL of saliva, color change first occurred in one of the positive control solutions at 30 minutes, so the 18S rRNA sequence amplified between 20 to 30 minutes (Table 6). Positive controls with 10^5^ copies of the 18S sequence/μL of saliva showed inconsistent amplification because some of the positive control samples stayed pink. Only one of the positive controls with saliva turned yellow before 40 minutes (Table 7).

**Table 6.** Amplification results with 10^6^ copies/µL 18S rRNA resuspended in saliva. Every 10 minutes in a vacuum flask filled with water at 65° C, the color of the solution in each reaction tube is scored as P (no amplification), O (some amplification), or Y (full amplification).

| Sample | 0 min | 10 min | 20 min | 30 min | 40 min | 50 min |
| --- | --- | --- | --- | --- | --- | --- |
| PT-1 (no saliva) | P | P | P | O | Y | Y |
| PT-2 | P | P | P | O | Y | Y |
| PT-3 | P | P | P | O | Y | Y |
| PT-4 | P | P | P | O | Y | Y |
| NT-1 (no saliva) | P | P | P | P | P | P |
| NT-2 | P | P | P | P | P | P |
| NT-3 | P | P | P | P | P | P |
| NT-4 | P | P | P | P | P | P |

**Table 7.** Amplification results with 10^5^ copies/µL 18S rRNA resuspended in saliva. Every 10 minutes in a vacuum flask filled with water at 65° C, the color of the solution in each reaction tube is scored as P (no amplification), O (some amplification), or Y (full amplification).

| Sample | 0 min | 10 min | 20 min | 30 min | 40 min | 50 min |
| --- | --- | --- | --- | --- | --- | --- |
| PT-1 (no saliva) | P | P | P | P | O | Y |
| PT-2 | P | P | P | P | O | O |
| PT-3 | P | P | P | P | P | O |
| PT-4 | P | P | P | P | P | P |
| NT-1 (no saliva) | P | P | P | P | P | P |
| NT-2 | P | P | P | P | P | P |
| NT-3 | P | P | P | P | P | P |
| NT-4 | P | P | P | P | P | P |

## 4. DISCUSSION

### 4.1 LAMP Results for Negative Controls with 10^3^ Copies of the 18S rRNA Sequence/μL of Molecular H_2_O

For negative control LAMP testing with the Han primer set, none of the reaction tubes changed color, which indicates that the primers did not falsely bind and indicate amplification without the presence of the 18S sequence throughout the 60-minute testing window. These results demonstrate that the Han primers may be suitable for detecting the 18S sequence and differentiating tubes with the 18S sequence present from those without, as it demonstrated a lack of amplification without the 18S sequence.

### 4.2 LAMP Results for Positive Controls with 10^3^ Copies of the 18S rRNA Sequence/μL of Molecular H_2_O

For LAMP testing with molecular H_2_O, the 18S sequence was amplified, thus the solutions changed color between 20 to 30 minutes in the positive controls, and the negative controls changed color between 40 to 50 minutes. Given the 20-minute time difference between the color changes in the positive and negative controls, the LAMP diagnostic was able to distinguish between samples with and without the 18S sequence. While the negative controls did show color change, as opposed to ideally staying pink throughout the testing window, it is important to note that this is not necessarily due to 18S rRNA detection. 50 minutes into a LAMP reaction, primer self-dimers may start to form through intermolecular interactions (Meagher et al. 2018). At this stage of the reaction, where six distinct primers are utilized, there is a significant likelihood of primer self-dimer formation. As a result, LAMP data beyond this point is typically considered unreliable and often disregarded.

As no color change was observed in one of the positive controls with 10^2^ copies of the 18S sequence/μL of molecular H_2_O during the LAMP reaction, the threshold for the Han primers in molecular water is 10^3^ copies of the 18S sequence/μL of molecular H_2_O. This indicates that the minimum 18S sequence concentration that the Han primers can detect in molecular water is 10^3^ copies of the 18S sequence/μL of molecular H_2_O.

### 4.3 LAMP Results for Positive Controls with 10^6^ Copies of the 18S rRNA Sequence/μL of Saliva

For LAMP testing with saliva, the 18S sequence was amplified, thus, the solutions changed color between 20 to 30 minutes in the positive controls, and the negative controls did not change color at all over the 50 minutes of the LAMP reaction. As the positive controls changed color while the negative controls did not, the LAMP diagnostic was able to distinguish between saliva samples with and without the 18S sequence.

As no color change was observed in one of the positive controls with 10^5^ copies of the 18S sequence/μL of saliva during the LAMP reaction, the threshold for the Han primers in molecular water is 10^6^ copies of the 18S sequence/μL of saliva. This indicates that the minimum 18S sequence concentration that the Han primers can detect in saliva is 10^6^ copies of the 18S sequence/μL of saliva.

### 4.4 Conclusions and Future Implications

Based on these results, LAMP is a cost and time-effective alternative for malaria testing. With LAMP, the Han primers detected a threshold concentration of 10^3^ copies of the *P. vivax* 18S sequence/μL of molecular water and 10^6^ copies of the *P. vivax* 18S sequence/μL of saliva. The *P. vivax* 18S sequence is amplified in the positive controls with both molecular water and saliva at 30 to 40 minutes into the LAMP reaction, which is quicker than the time required for qPCR and blood microscopy to detect *P. vivax*. Additionally, a vacuum flask filled with water at 65° C can be substituted for the water bath, as it does not result in any changes in the LAMP results for the positive and negative controls.

As the implications of this project are aimed at real-world applications for malaria testing in low-resource settings where malaria is pervasive, there must be further experimentation to test the effectiveness of LAMP in detecting other malaria strains, such as *P. falciparum*. Additionally, the remote LAMP diagnostic can be further optimized to be more accessible and user-friendly in these settings with features such as a self-heating system and the substitution of liquid reagents for dried reagents, such as lyophilized LAMP master mix. Ultimately, the results of this study demonstrate the high efficacy of LAMP in detecting *P. vivax* in saliva using cost-effective equipment. This finding has significant implications for developing more accessible diagnostic tests for *P. vivax* malaria. By providing a reliable and affordable diagnostic tool, this approach has the potential to revolutionize malaria diagnostics and significantly improve public health outcomes.

## Data Availability

All data produced in the present work are contained in the manuscript.

## 5. ACKNOWLEDGMENTS

This work was made possible thanks to donations from Bob and Cathy O’Rear and New England Biolabs. We would like to acknowledge support from the community of student researchers that make up the UT Austin CNS FRI DIY Diagnostic Research Lab (https://diystream.cns.utexas.edu/) and the UT Austin Ellington Lab (https://ellingtonlab.org/).

